# Comparing phrenic nerve stimulation using three rapid coils: implications for mechanical ventilation

**DOI:** 10.1101/2022.03.29.22272862

**Authors:** Kyle G. Boyle, Philipp A. Eichenberger, Patrick Schön, Christina M. Spengler

**Affiliations:** Exercise Physiology Lab, Institute of Human Movement Sciences and Sports, ETH Zurich, Zurich, Switzerland; Zurich Center for Integrative Human Physiology (ZIHP), University of Zurich, Zurich, Switzerland

## Abstract

**Rationale:** Rapid magnetic stimulation (RMS) of the phrenic nerves may serve to attenuate diaphragm atrophy during mechanical ventilation. With different coil shapes and stimulation location, inspiratory responses and side-effects may differ.

**Objective:** To compare the inspiratory and sensory responses of three different RMS-coils either used bilaterally on the neck or on the chest, and to determine if ventilation over 10min can be achieved without muscle fatigue and coils overheating.

**Methods:** Healthy participants underwent bilateral anterior 1-s RMS on the neck (RMS_BAMPS_) (n=14) with three different pairs of magnetic coils (parabolic, D-shape, butterfly) at 15, 20, 25 and 30Hz stimulator-frequency and 20% stimulator-output with +10% increments. The D-shape coil with individual optimal stimulation settings was then used to ventilate participants (n=11) for up to 10min. Anterior RMS on the chest (RMS_aMS_) (n=8) was conducted on an optional visit. Airflow was assessed via pneumotach and transdiaphragmatic pressure via esophageal and gastric balloon catheters. Perception of air hunger, pain, discomfort and paresthesia were measured with a numerical scale.

**Main results:** Inspiration was induced via RMS_BAMPS_ in 86% of participants with all coils and via RMS_aMS_ in only one participant with the parabolic coil. All coils produced similar inspiratory and sensory responses during RMS_BAMPS_ with the butterfly coil needing higher stimulator-output, which resulted in significantly larger discomfort ratings at maximal inspiratory responses. Ten of 11 participants achieved 10min of ventilation without decreases in minute ventilation (15.7±4.6L/min).

**Conclusions:** RMS_BAMPS_ was more effective than RMS_aMS,_ and could temporarily ventilate humans seemingly without development of muscular fatigue.

## Introduction

The use of mechanical ventilation (MV) to replace spontaneous breathing is the gold standard during respiratory failure. However, the unphysiological positive pressure and prolonged diaphragmatic inactivity during MV can induce lung injury and diaphragm atrophy within 18h (1). Atrophy is associated with intramuscular changes at fiber and cellular levels (2-4), leading to a reduced ability to generate force and thus prolonged weaning time (5), increasing the risk of further damage and pulmonary complications.

One potential method to reduce ventilator-associated diaphragm atrophy is to activate the diaphragm via nerve-stimulation. Preliminary results in animals (6, 7) have shown that phrenic nerve stimulation (PNS) reduces MV-induced diaphragm atrophy. Furthermore, diaphragm pacing via PNS in spinal cord injured subjects can replace MV to induce resting breathing and preserve diaphragm function (8). Intravenously inserted stimulation catheters have also been tested (9) and were recently shown to attenuate the decrease in inspiratory muscle strength, but to not decrease weaning time in difficult-to-wean patients (10). However, any catheter poses a risk for infection and thus, non-invasive external PNS may serve as a favorable alternative.

Magnetic stimulation is one method for non-invasive PNS via a variety of coils and stimulation locations such as over the cervical spine (CMS) (11), bilaterally and anterolaterally on the neck (12), or anteriorly on the chest (aMS) (13). Two research groups have explored the use of rapid magnetic stimulation (RMS) in the context of diaphragm stimulation (14, 15), and this technique was recently shown to not negatively interact with intensive care unit (ICU) equipment (16). RMS may therefore serve as an ideal candidate for non-invasive treatment. Sander *et al*. (14) showed that rapid bilateral anterior magnetic phrenic nerve stimulation (RMSBAMPS) can produce strong enough diaphragmatic contractions to induce short-term ventilation in healthy subjects for no longer than 5min due to “technical prerequisites.” Adler *et al*. (15) sought to determine the optimal combination of stimulation-frequency and stimulator-output for tolerable rapid CMS (RMS_CMS_) in healthy humans, but they were unable to induce inspiratory flow despite sufficient diaphragm contraction.

With new developments of coils and cooling techniques, the present study aimed to compare inspiratory and sensory responses to RMS_BAMPS_ when using different combinations of stimulation-frequency and stimulator-output with three differently-shaped coils, and to investigate whether a continuous series of RMS_BAMPS_ could sustain ventilation at a constant level for 10min. Additionally, rapid anterior stimulation on the chest (RMS_aMS_) was tested, as this location may offer better nerve access.

## Methods

### Ethical approval

The study was approved by the Cantonal Ethics Committee of Zurich (Project ID 2019-01990) and registered on clinicaltrials.gov (NCT04176744). The study conformed with the *Declaration of Helsinki*.

### Experimental design

The study took place over 2-3 study visits. Participants abstained from intense exercise 48h before, and from any type of exercise 24h before each study visit, and from consumption of alcohol and caffeinated food or drinks on study days prior to testing. During visit 1, participants underwent lung function and respiratory muscle testing followed by a single-train RMS_BAMPS_ protocol consisting of 1-s RMS trains of the phrenic nerves. The single-train RMS_BAMPS_ protocol was conducted with three commercially available coils using various combinations of stimulator-output (% of maximum) and stimulation-frequency. During visit 2, participants had their body composition assessed prior to undergoing a series of consecutive trains of RMS_BAMPS_ to induce ventilation in 1-min blocks, using the cooled D-shape coils, for a maximum of 10min. During the optional visit 3, participants underwent the same protocol as during visit 1, except that RMS_BAMPS_ was replaced by RMS_aMS_ performed on the chest. Cardiorespiratory variables, respiratory muscle activation, subjective perceptions and measures of side-effects were monitored throughout all visits.

### Participants

Of the 15 participants originally recruited for the study, one participant withdrew due to an initial misunderstanding of study requirements. Thus, data of 14 participants (9M:5F) are presented. Due to the Covid-19 pandemic, only five participants (4M:1F) returned for the optional study visit and three additional participants (3F) were recruited at the end of the study to only undergo the optional visit 3 (RMS_aMS_). Participant characteristics including anthropometrics, body composition, lung function and respiratory muscle strength are given in Table 1.

**Table 1.** Participant Characteristics.

|  | Initial Recruitment | Secondary Recruitment |
| --- | --- | --- |
| **Anthropometrics** |  |  |
| Sex | 9M:5F | 0M:3F |
| Age, years | 26 ± 5 | 28 ± 4 |
| Height, cm | 176 ± 9 | 169 ± 4 |
| Body weight, kg | 74 ± 10 | 58 ± 6 |
| Body fat, kg | 18 ± 5 | 15 ± 3 |
| Body fat, % | 26 ± 6 | 29 ± 3 |
| BMI, kg/m<sup>2</sup> | 24 ± 2 | 20 ± 1 |
| **Spirometry** |  |  |
| FVC, L | 5.4 ± 1.2 | 4.6 ± 0.7 |
| FVC, % predicted | 106 ± 10 | 110 ± 11 |
| FEV<sub>1</sub>, L | 4.4 ± 0.8 | 3.7 ± 0.6 |
| FEV<sub>1</sub>, % predicted | 102 ± 9 | 106 ± 12 |
| FEV<sub>1</sub>/FVC, % | 81 ± 7 | 81 ± 5 |
| Maximal voluntary ventilation, L/min | 164 ± 42 | 129 ± 22 |
| Maximal voluntary ventilation, % predicted | 106 ± 16 | 99 ± 3 |
| **Maximal volitional pressure generation** |  |  |
| Maximal inspiratory mouth pressure, cmH<sub>2</sub>O | 117 ± 17 | 104 ± 19 |
| Maximal inspiratory mouth pressure, % predicted | 121 ± 21 | 125 ± 37 |
| Maximal expiratory mouth pressure, cmH<sub>2</sub>O | 155 ± 36 | 148 ± 53 |
| Maximal expiratory mouth pressure, % predicted | 121 ± 30 | 150 ± 53 |

*Definition of abbreviations:* BMI = body mass index; FVC = forced vital capacity; FEV<sub>1</sub> = forced expiratory volume in one second. Values are mean±SD. Predicted FVC and FEV<sub>1</sub> values were obtained from Quanjer et al. (2012) (18) and predicted maximal inspiratory and expiratory values were obtained from Wilson et al. (1984) (19). Predicted maximal voluntary ventilation was obtained by multiplying FEV<sub>1</sub> by 35. Secondary recruitment represents the participants who were recruited for the optional third visit only (rapid magnetic stimulation on the chest).

### Lung function, respiratory muscle strength, body composition

In all participants standard lung function, respiratory muscle strength, and body composition was assessed. Lung function tests were performed using a commercially available testing system and body box (Quark PFT & Q-Box, Cosmed, Rome, Italy) according to current guidelines (17). Respiratory muscle strength was assessed via maximal inspiratory and expiratory maneuvers (from residual volume and total lung capacity, respectively) alternating every three maneuvers using a respiratory pressure meter (RP Check, MD Diagnostics LTD., Kent, United Kingdom). Percentage of predicted values were calculated for each participant using specific reference equations for lung function (18) and respiratory muscle strength (19). Body composition was measured via a dual-energy X-ray absorptiometry (DXA) scan (lunar iDXA densitometer, GE Healthcare, Madison, WI, United States).

### Cardiorespiratory response to RMS

Participants were instrumented with a mouthpiece connected to a calibrated pneumotachometer (Series 3813, Hans Rudolph, Shawnee, KS, USA) for continuous measurement of flow. End-tidal partial pressure of carbon dioxide (P_ET_CO_2_) and peripheral blood oxygen saturation (S_P_O_2_) were measured using a patient monitor (Cardiocap/5, Datex-Ohmeda, Madison, WI, USA). On all visits, participants had the option of inserting two balloon catheters (Adult Esophageal Balloon Catheters 47-9005, Cooper Surgical, Trumbull, CT, USA) that were connected to calibrated differential pressure transducers (DP45, Validyne Engineering, Northbridge, CA, USA) to record transdiaphragmatic pressure (P_di_). Briefly, both balloon catheters were inserted through the nares, following numbing with local anesthetic spray (Xylocain Spray 10%, Aspen Pharma Schweiz GmbH, Baar, Switzerland), and both balloons were first placed in the stomach. One balloon remained in the stomach to measure gastric pressure (P_ga_). The second balloon was withdrawn in 1cm increments until a negative deflection was detected during a sniff maneuver, followed by the balloon being withdrawn an additional 10cm to ensure it was completely removed from the stomach in order to measure esophageal pressure (P_es_). Participants were instructed to execute a Valsalva maneuver to empty both the gastric and esophageal balloons which were then filled with 2ml and 1ml of air, respectively. Final placement was adjusted, if needed, so that end-expiratory pressure during tidal breathing resulted in a P_es_ of 0cmH_2_O. Both balloons were then secured to the nose with tape to ensure the same position throughout the experiments. P_di_ was calculated as the difference between P_ga_ and P_es_. All participants elected to attempt catheter insertion but only 8 of the 14 tolerated the procedure. To determine if less invasive measures of diaphragm contraction would correlate with V_T_ and P_di_, participants were also equipped with two respiratory belts (TN1132/ST, ADInstruments, Dunedin, New Zealand). One belt was placed over the naval, while one was placed along the nipple line in men and the highest possible position below the breasts in women to measure changes in abdominal and chest circumference, respectively (Δ_Abdominal_ and Δ_Chest_). Finally, cardiovascular changes in response to RMS were monitored with a simple 3-lead electrocardiogram connected to a bioamplifier (PowerLab 15T, Dunedin, New Zealand) and a plethysmographic finger cuff (Nexfin, Edwards Lifesciences, Amsterdam, Netherlands) for continuous measurement of heart rate (HR) and blood pressure (BP), respectively.

### Side-effects in response to RMS

Participants rated their perception of pain, discomfort and paresthesia in response to each RMS setting by pointing to a visual scale ranging from 0-to-10points. 0 was anchored as “none” while 10 was anchored as “maximal” (the maximum that one could imagine). Participants were also asked specifically whether they felt dental pain. During the continuous RMS_BAMPS_-ventilation trial on visit 2, the perception of air hunger was also evaluated via the same 0-to-10point visual scale. Change in galvanic skin response (GSR), a surrogate measure for changes in one’s emotional response, was measured on the fingers using a commercially available GSR system (MLT118F GSR Electrodes & FE116 GSP Amp, ADInstruments, Dunedin, New Zealand). Lastly, the presence of upper air way collapse during RMS was quantified by counting the instances of an increase in P_di_ or Δ_Abdominal_ without concomitant flow as seen in Figure 1B.

**Figure 1.**
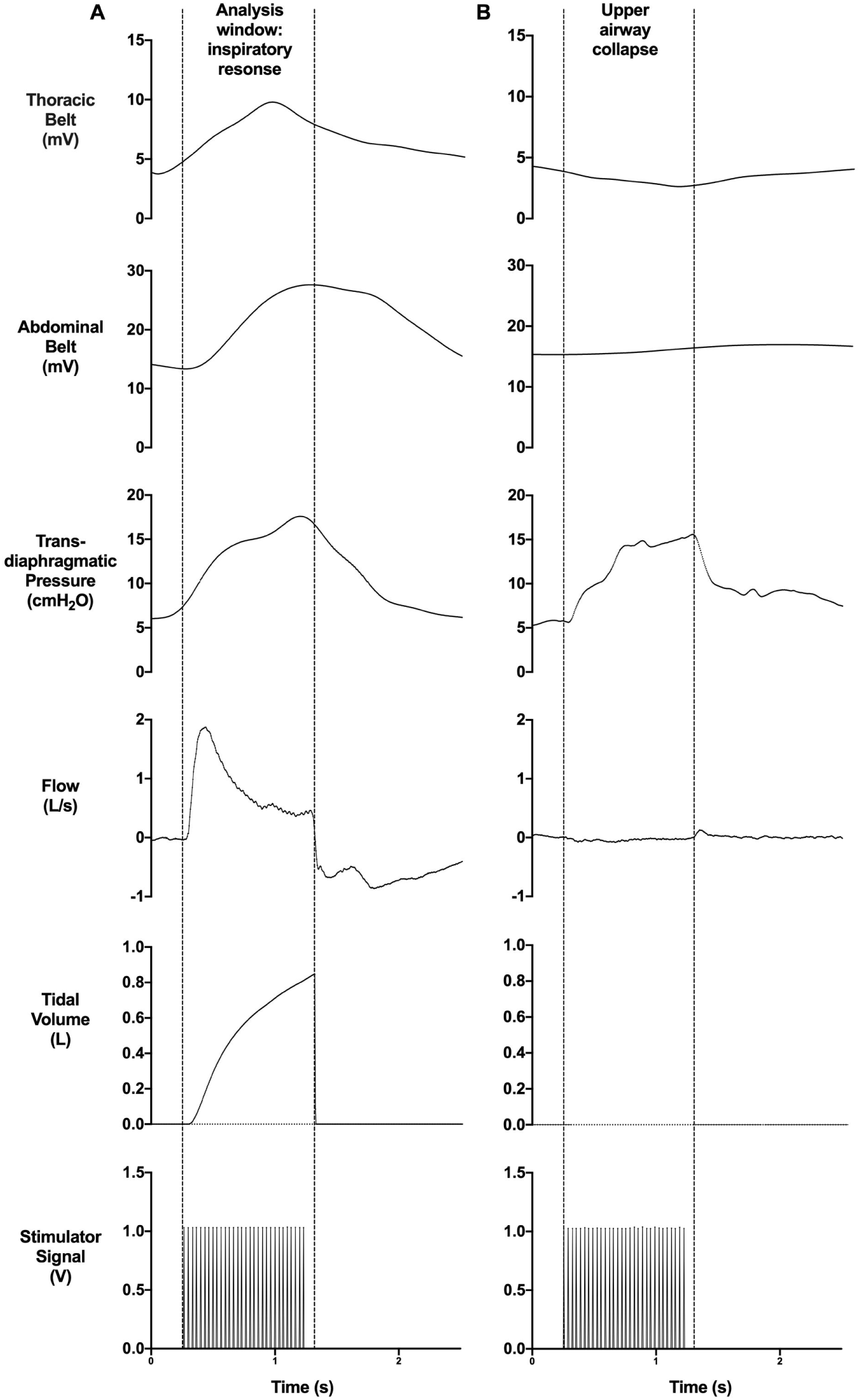
Example of the data analysis window during an inspiratory response and of an upper airway collapse induced by rapid magnetic stimulation. A: Thoracic excursion, abdominal excursion, transdiaphragmatic pressure, flow and tidal volume in response to rapid magnetic stimulation of the phrenic nerves with the glottis open. Dashed lines between the start of the stimulator signal and the end of inspiration represent the analysis window in which the mean and/or peak values were used in the analysis of select variables. B: Example of an upper airway collapse during rapid magnetic stimulation of the phrenic nerves represented by an increase in transdiaphragmatic pressure without the presence of flow. Both A and B are from rapid magnetic phrenic nerve stimulation performed bilaterally and anteriorly on the neck using the D-shape (30Hz stimulation-frequency, 20% stimulator-output) and butterfly (30Hz stimulation-frequency, 30% stimulator-output) coils, respectively. Data within figure has been smoothed.

### Single-train RMS_BAMPS_ *(visit 1)*

Single-train RMS_BAMPS_ was conducted using two commercially available magnetic stimulators (MagPro X100, MagVenture, Farum, Denmark) and 3 pairs of differently-shaped magnetic coils: a parabolic-shaped coil (MMC-90, MagVenture, Farum, Denmark [max dB/dt=30kT/s, 280µs]), a butterfly-shaped coil (Cool-B65, MagVenture, Farum, Denmark [max dB/dt=36kT/s, 290µs]), and a D-shaped coil (Cool-D50, MagVenture, Farum, Denmark [max dB/dt=27kT/s, 248µs]). The butterfly and D-shaped coils were also equipped with an active cooling unit (Coil cooler unit + high performance option, MagVenture, Farum, Denmark). All RMS_BAMPS_ took place with the participant laying in a hospital bed that was tilted upright so that the torso was raised 30°. Participants’ heads were positioned within a vacuum cushion (Vacuform® 2.0 vacuum pillow 30×40cm, Synmedic AG, Zurich, Switzerland) so that their necks were slightly extended. Positions of anatomical landmarks were recorded to ensure that all stimulations occurred with the participant in the same head and body position, as well as to ensure that the body position could be accurately repositioned on subsequent visits (for details also see below).

The single-train RMS_BAMPS_ protocol began with 3min of resting breathing to quantify baseline measures and subsequently the placement of the first two coil anterolaterally on the neck in the position that yielded the highest response to stimulation. The coils were initially placed in the position that yielded the highest P_di_ in response to a single bilateral stimulation (P_di,tw_) or - in participants who did not tolerate catheter insertions - the highest tidal volume (V_T_) in response to a 1-s train of RMS_BAMPS_ at 20Hz and 20-40% of stimulator-output, depending on participant tolerance. Final adjustments of coil positions were made to minimize any excessive movement of the arms and head during RMS_BAMPS._ The final position was secured with custom-made lever arms and recorded using the Brainsight® neuro-navigation system (BrainSight® TMS, Rogue Research Inc., Montreal, Canada) with reflective markers placed on the coils and on the participant via custom made glasses. For a figure of the setup, please contact the corresponding author. A maximum of 1h was used to place the coils.

Following placement of the coils, trains of RMS_BAMPS_ were systematically conducted at 15, 20, 25 and 30Hz in a randomized order. For each frequency, the initial stimulator-output was set at 20% of maximum and increased in 10% increments until the participant no longer tolerated the increase. These 1-s trains of RMS_BAMPS_ occurred at the end of a passive expiration with the glottis open and upon signal of the participant. All stimulation-frequency and stimulator-output combinations were applied at least twice unless participants indicated they could not tolerate a second train at that setting. In the event of data contamination (ie. swallow, upper airway collapse, improper timing, etc.), additional trains of RMS_BAMPS_ were conducted at that setting on tester’s discretion. Following RMS_BAMPS_ with the four tested frequencies and all tolerable stimulator-outputs, the protocol was repeated starting with coil placement of the second pair of coils and after completion of that protocol, with the third pair. Coils were tested in a randomized order. Lastly, if a participant did not show any flow response, visit 1 was repeated on another day, at least 24h apart of the first visit 1. If no flow could be initiated on the second day, these participants were considered as non-responders.

### Continuous RMS_BAMPS_-ventilation (visit 2)

Participants in whom flow was induced via RMS_BAMPS_ underwent thorough familiarization with continuous RMS_BAMPS_-ventilation. During this familiarization, the optimal stimulation-frequency and stimulator-output combination of visit 1 - using the pair of cooled coils that had yielded the largest VT at a submaximal stimulator-output (always the D-shape coils) - was tested at a respiratory frequency selected initially to match resting ventilation. If needed, adjustments to stimulator-output (±5% increments), stimulation duration (+1ms increments) and respiratory frequency (+1breath-per-min increments) were made in attempt to optimally balance subjectively and objectively sufficient ventilation as well as suppression of participants’ natural drive to breathe, while keeping their perception of air hunger, pain, discomfort and paresthesia tolerable throughout. As such, participants’ verbal feedback was sought and taken into account to optimize their ability to complete the ten 1-min blocks of continuous RMS_BAMPS_-ventilation, as well as to reduce their urge for spontaneous breathing.

After familiarization, a 3-min resting breathing period was recorded followed by 1min of ventilation. Between each train of RMS_BAMPS_, participants passively expired and were instructed to not initiate the next inspiration. The flow signal was continuously monitored to ensure participants did not initiate breaths themselves. After each 1-min stimulation block, participants were asked to rate their perception of air hunger, pain, discomfort, and paresthesia, and whether they could tolerate a further minute of continuous RMS_BAMPS_. This break lasted the minimum amount of time to collect the sensory ratings. This procedure was repeated until ten 1-min blocks were completed or until participant cessation.

### Single-train RMS_aMS_ *(visit 3)*

The optional study visit 3 replicated the RMS_BAMPS_ protocol from visit 1 (n=5) with RMS_aMS_ conducted on the chest. Coils were tested in two positions: 1) two coils placed bilaterally on either side of the chest; and 2) a single coil placed in the middle of the chest in attempt to stimulate both phrenic nerves simultaneously. A maximum of 1h was spent to optimize the position. The 3 additionally recruited subjects performed a DXA scan, lung function and respiratory muscle strength tests and the same RMS_aMS_ protocol.

### Data acquisition and analysis

All physiological measurements were converted from analogue to digital with two 16-channel data acquisition systems (PowerLab 16/35, ADInstruments, Dunedin, New Zealand) and collected using LabChart Software (Version 8, ADInstruments, Dunedin, New Zealand) with a sampling frequency of 2,000Hz. VT was calculated by taking the integral of the flow measurement which was then BTPS corrected. Within a data analysis window that started at the beginning of RMS and ended at the absence of inspiratory flow (Figure 1A), V_T_, P_di,mean_ and P_di,peak_, as well as mean Δ_Abdominal_ and Δ_Chest_ were calculated. Peak ΔGSR was analyzed outside of this window due to the latency of this signal. All responses with the same RMS setting that did not induce total upper airway collapse and where pairs of stimulations were available, were included in the analysis of inspiratory variables and averaged. Stimulations that induced total upper airway collapse were not included in the analysis of inspiratory variables, but were still quantified to determine the prevalence of this side-effect. Responses during the continuous RMS_BAMPS_-ventilation trial were analyzed breath by breath in the same manner as the single-train RMS and averaged over 1 min, not including the short breaks. In addition, an average of all 1-min blocks was calculated to get a total overview of the RMS_BAMPS_-ventilation trial. The tension-time-index of the diaphragm (TTI_di,mean_) was calculated as the average pressure the diaphragm produced during inspiration compared to the maximum a participant could achieve during a maximal maneuver (P_di,mean_/P_di,max_) multiplied by the duty cycle (inspiratory duration/breath cycle). Non-responders were not included in the analysis.

A number of factors contributed to an unequal sample size within and between coils at select RMS settings (Figure 3A) and between visits (Figure 2). First, not all participants were able to tolerate the same maximal RMS settings within and between coils. Second, not all participants were equipped with balloon catheters during each visit. Third, one participant was not able to undergo single-train RMS_BAMPS_ with the butterfly coil due to too much facial paresthesia. Fourth, one participant was unable to return for visit 2 due to the COVID-19 pandemic. Finally, only five of 12 participants returned for the optional visit 3, so three additional participants were recruited to undergo visit 3 only. Therefore, the number of people included in the analysis is summarized in Figure 2.

**Figure 2.**
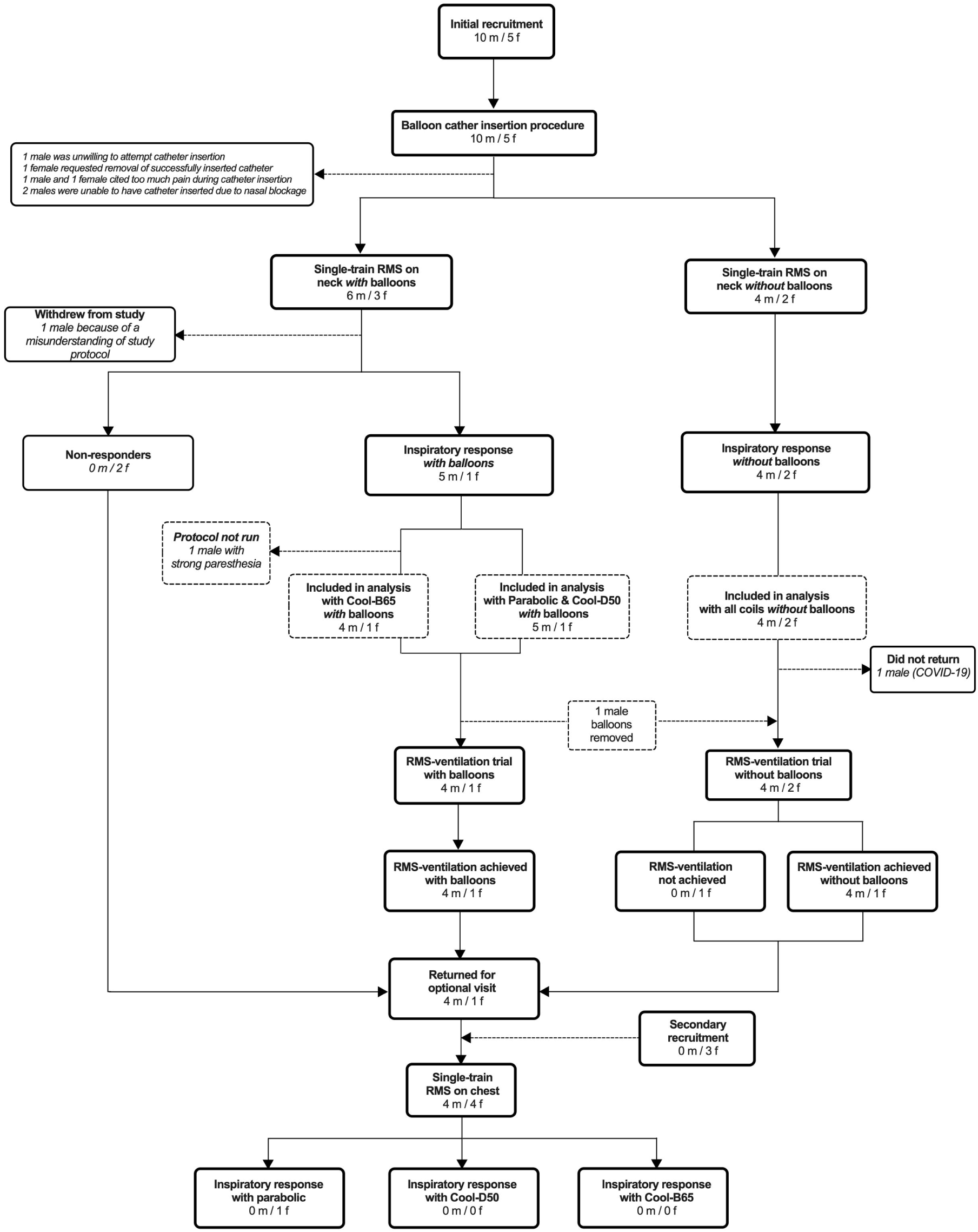
Summary of participant recruitment and analysis. *Definition of abbreviations*: RMS = rapid magnetic stimulation. Fifteen participants were initially recruited, and one participant withdrew. As such, 14 participants were included in the analysis for single-train RMS on the neck (RMS_BAMPS_), which occurred on visit 1. Of those 14 participants, 8 participants were equipped with balloon catheters and two of those were non-responders. One participant with balloons did not undergo single-train RMS_BAMPS_ with the butterfly coil due to facial paresthesia. Therefore, 12 participants (six with balloons) could be analyzed with the parabolic and D-shape coil, while 11 participants (five with balloons) could be analyzed with the butterfly coil. One participant (without balloons) was unable to return for the RMS_BAMPS_-ventilation trial (visit 2) due to the COVID-19 pandemic, while one participant who was equipped with balloons on visit 1 declined to undergo balloon catheter insertion on visit 2. One additional participant (without balloons) was unable to be ventilated with continuous RMS_BAMPS_. As such, ten participants (five with balloons) could be analyzed for RMS_BAMPS_-ventilation. Five participants elected to return for the optional visit in which single-train RMS was conducted on the chest, and three additional participants were recruited to undergo this visit only. Only one participant (without balloons) showed an inspiratory response to RMS on the chest with the parabolic coil only and could be analyzed.

**Figure 3.**
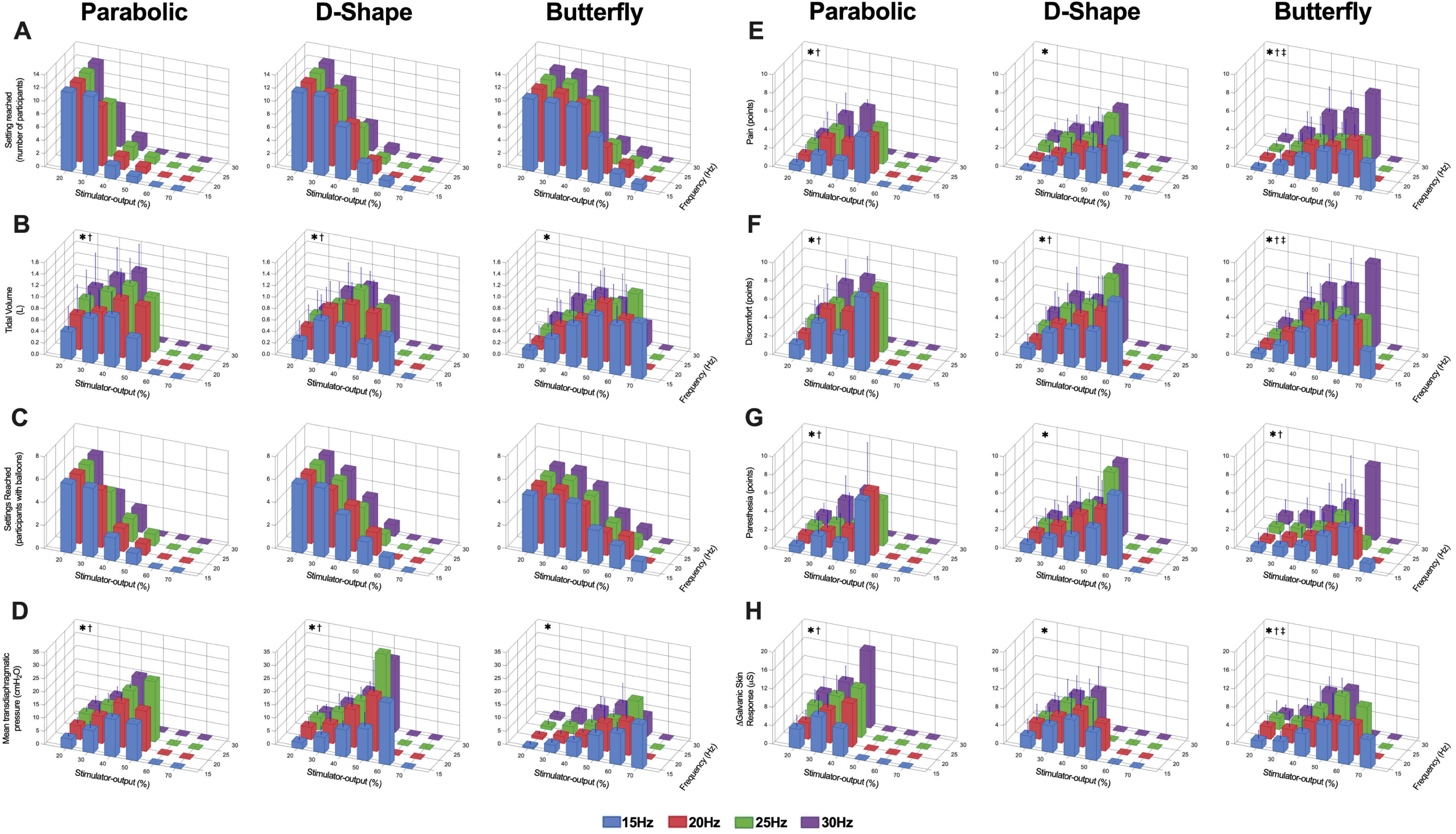
Inspiratory and side-effect responses to rapid magnetic stimulation bilaterally on the neck (RMS_BAMPS_) with different stimulator-output and stimulation-frequency combinations. A: Number of subjects that showed an inspiratory response to RMS_BAMPS_ that reached each tested stimulation setting; one participant did not undergo RMS_BAMPS_ with the butterfly coil. B. Tidal volume responses to RMS_BAMPS_ (n=panel A). C: Number of subjects who were equipped with balloon catheters that showed an inspiratory response to RMS_BAMPS_ and reached each tested stimulation setting. D: Mean transdiaphragmatic pressure responses to RMS_BAMPS_ (n=panel C). E-G: Pain, discomfort and paresthesia in response to RMS_BAMPS_ (n=panel A). H: Change in galvanic skin response in response to RMS_BAMPS_ (n at 20%=10 due to technical difficulties). *, main effect of stimulator-output; ^†^, main effect of stimulation-frequency; ^‡^, interaction effect (P<0.05). Values in panels A and C are absolute count, while all other values are mean+SD.

### Statistics

Due to the unequal sample size with each RMS setting, a mixed effects model two-way ANOVA with repeated measures was used to determine the effect of stimulation-frequency and stimulator-output on V_T_, P_di_, sensory ratings and ΔGSR within each coil. Coils were compared between each other at select V_T_ using a mixed effects model one-way ANOVA with repeated measures. Tukey’s post hoc test was used in the event of a significant effect to determine which specific coils differed. A Pearson’s correlation coefficient was used to determine within-participant relationships between ΔA_bdominal_ and Δ_Chest_ with both V_T_ and P_di,mean_; while a Spearman correlation coefficient determined within-participant relationships between ΔGSR and all sensory ratings during single-train RMS_BAMPS_. A repeated measures correlation was used to determine overall relationships between the same variables. Paired *t*-tests were used to compare mean continuous RMS_BAMPS_-ventilation trial responses with resting breathing, while a linear regression was used to determine if cardiorespiratory or sensory values significantly increased or decreased over time. All statistical analyses were conducted using Prism (v8.3.1, GraphPad Software, San Diego, CA, USA) except for the repeated measures correlation that was conducted using the RStudio’s (v1.4.1106, Boston, MA, USA) “rmcorr” package. All values are expressed as mean±SD unless otherwise stated. *P*-values less than 0.05 were considered statistically significant.

## Results

### Effect of stimulation-frequency and stimulator-output on inspiratory responses

Inspiration in response to single-train RMS_BAMPS_ was induced in 12 of 14 participants. Both non-responders were female (with P_di_ measurement) and showed the same non-response on two visits. In general, increasing both stimulator-output and stimulation-frequency resulted in an increase in V_T_ (n=12) and P_di_ (n=6) as displayed in Figure 3B&D, with larger changes with stimulator-output compared to frequency increases. Increased stimulator-output had a significant effect on V_T_ with all coils (all *P*≤0.003); while an effect of frequency was only present with parabolic (*P*=0.023) and D-shape (*P*=0.041) coils, without an interaction effect. There was a significant effect of stimulator-output on P_di,mean_ (all *P*≤0.005) and P_di,peak_ (all *P*≤0.002) with all coils, while only the D-shape and parabolic coils had an effect of frequency with both variables (all *P*≤0.036), and no coil displayed an interaction effect. Both V_T_ and P_di,mean_ showed a stronger correlation with Δ_Abdominal_ than with Δ_Chest_ (Table 3).

### *Side-effects of RMS*_BAMPS_

In general, sensory responses increased more prominently with increases in stimulator-output than increases in frequency (Figure 3E-G). The largest stimulator-output tolerated in all stimulation-frequency and coil combinations was 20% (Figure 3A). For pain perception, all coils showed significant effects of stimulator-output (all *P*≤0.0002). The parabolic (*P*=0.012) and butterfly (*P*<0.0001) coil showed a significant effect of frequency, and the butterfly only showed an interaction (*P*=0.002). For discomfort, all coils had a significant effect of stimulator-output (all *P*<0.0001) and frequency (all *P*≤0.037), with a significant interaction (*P*=0.016) in the butterfly only. For paresthesia, all coils had an effect of stimulator-output (all *P*≤0.004), but an effect of frequency was only present with parabolic (*P*=0.011) and butterfly (*P*=0.005) coils, without interaction with any coil. One participant was unable to undergo RMS with the butterfly coil citing too much facial paresthesia.

Changes in GSR in response to single-train RMS_BAMPS_ are given in Figure 3H. Two participants were excluded from analysis due to technical difficulties. An effect of stimulator-output on ΔGSR was present in all coils (all *P*≤0.0005), an effect of frequency was only present in parabolic (*P*=0.010) and butterfly (*P*=0.0499) coils, while an interaction was only present with the butterfly (*P*=0.011). ΔGSR showed a moderate positive correlation with both pain (r=0.55, *P*<0.001) and discomfort (r=0.63, *P*<0.001), and a weak positive correlation with paresthesia (r=0.38, *P*<0.001) (Table 3).

All incidents of UAC and dental pain are shown in Table 2. UAC was a common side-effect most prevalent when stimulating at the lowest frequency (15Hz) and stimulator-output (20%). UAC was most likely to occur with the D-shape coil and least likely with the butterfly. The prevalence of dental pain (most prevalent with the butterfly coil) increased with increasing stimulator-output and occurred more often at 25 and 30Hz compared to 15 and 20Hz.

**Table 2.**
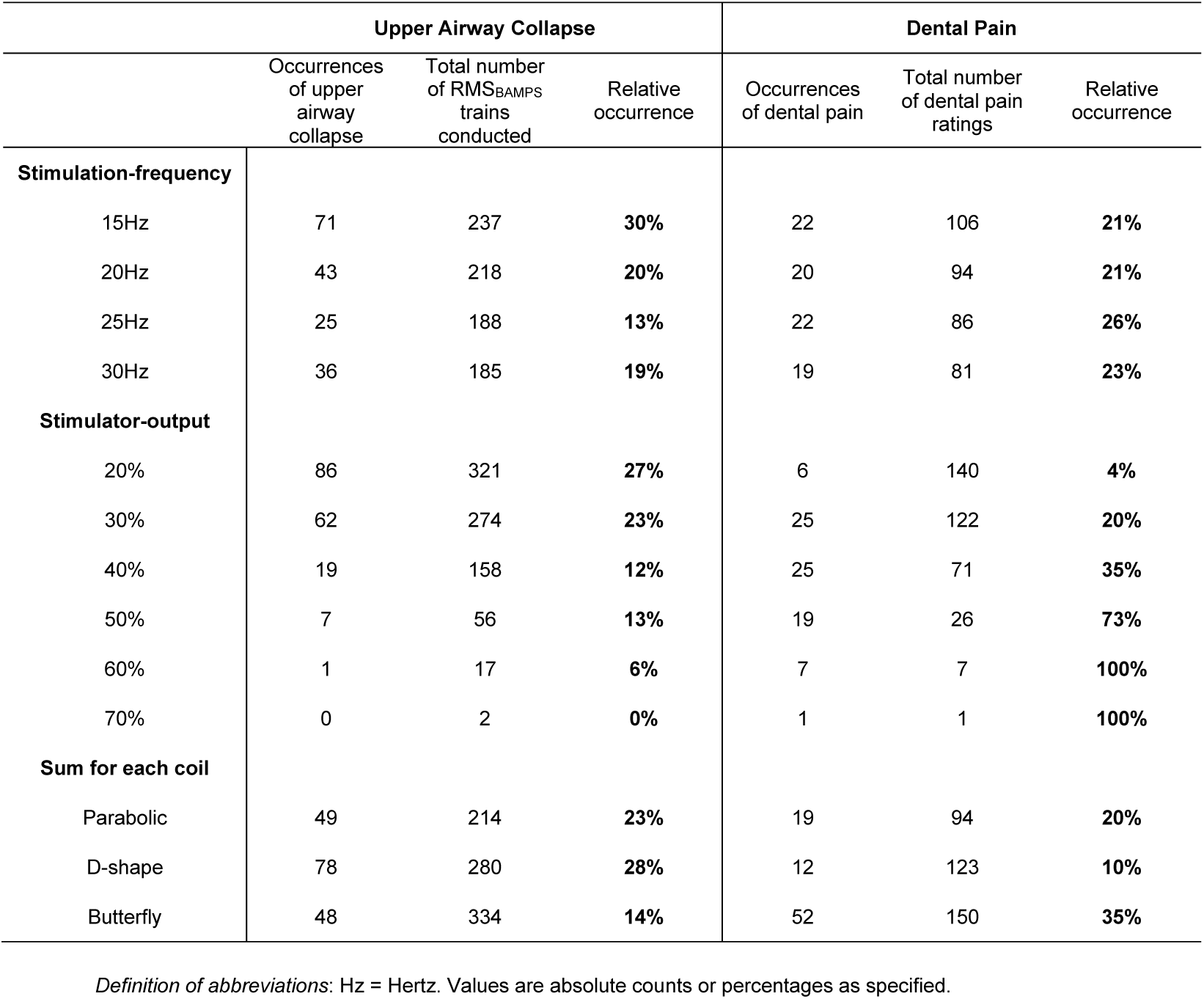
Prevalence of upper airway collapse and dental pain during single-train rapid magnetic stimulation bilaterally on the neck (RMS_BAMPS_).

### Between coil comparisons

For any given stimulation-frequency and stimulator-output combination, the parabolic and D-shape coils achieved larger V_T_ compared to the butterfly. With the butterfly, participants tolerated higher levels of stimulator-output achieving the same maximal V_T_ as with the other coils at the highest tolerated stimulator-output (all *P*>0.05), but with significantly larger pain (vs. parabolic +1.2±1.7points, *P*=0.046; vs. D-shape +1.5±1.9points, *P*=0.030) at 30Hz. The highest achieved V_T_ and the associated sensory ratings are presented in Figure 4. Maximal V_T_ did not differ between coils (parabolic 1.08±0.40L; D-shape 1.11±0.34L; butterfly 1.06±0.41L, *P*=0.804), but D-shape and butterfly coils needed more stimulator-output to achieve these volumes (parabolic=29±9%; D-shape=37±8%; butterfly=45±9%). Corresponding Pdi,mean in participants who were equipped with balloon catheters during these stimulations was 10.7±5.5 (parabolic), 14.4±9.7 (D-shape) and 8.8±5.3cmH_2_O (butterfly). During these maximal stimulations, there was a significant effect of coil on discomfort (*P*=0.047) with post-hoc tests revealing significantly higher ratings between parabolic and butterfly coils (4.9±2.0 vs. 6.5±2.3points, *P*=0.046), but no difference in pain nor paresthesia. When considering only stimulations that resulted in a V_T_ between 4-8ml/kg body weight (Figure 5), which reflects the target V_T_ during MV in patients with acute respiratory distress syndrome (ARDS) (20), sensory ratings were smaller compared to stimulations where maximal V_T_ was reached. Mean pain, discomfort and paresthesia ratings during those stimulations was 1.1±1.1, 2.5±1.9 and 0.8±1.1points (parabolic), 1.4±1.2, 2.3±1.6 and 1.5±1.6points (D-shape) and 2.0±2.1, 3.3±2.4 and 0.9±1.4points (butterfly).

**Figure 4.**
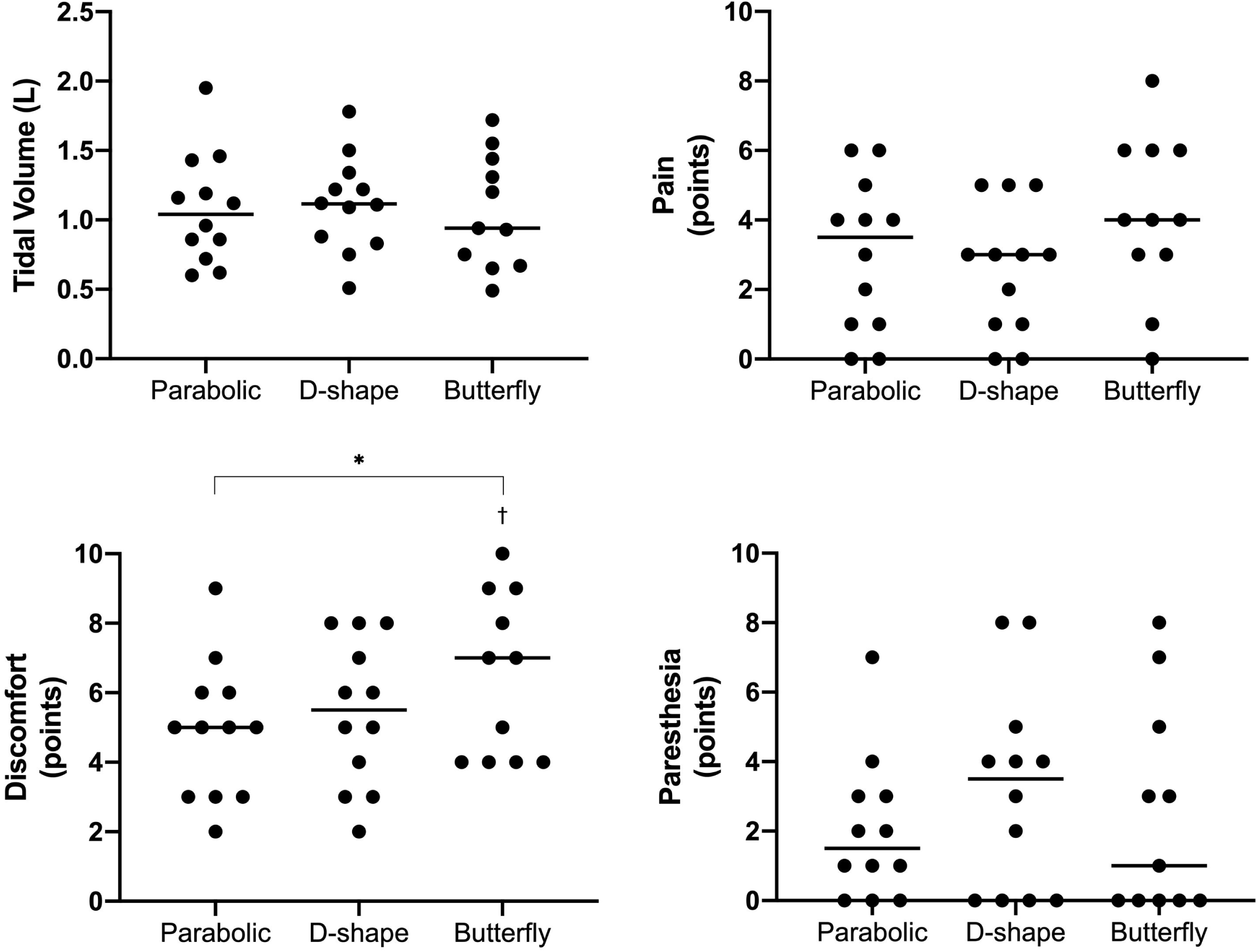
Highest tidal volume achieved in each coil in response to single-train rapid magnetic stimulation bilaterally on the neck (RMS_BAMPS_) and associated sensory ratings with each coil. Black horizontal line in all panels represent mean values, while black dots represent individual participant values. N=12 except for the butterfly coil data (n=11) as one participant did not undergo RMS_BAMPS_ with that coil. *, main effect of coil; ^†^, significantly different from parabolic (P<0.05).

**Figure 5.**
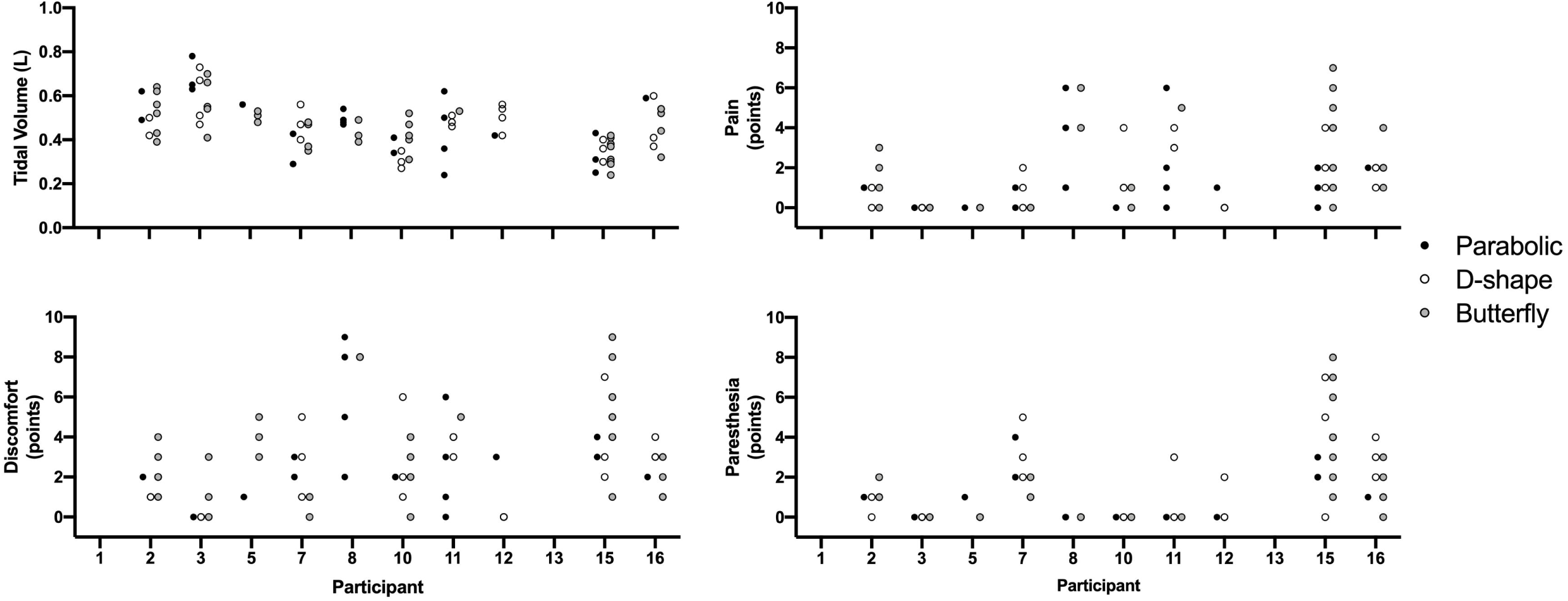
Tidal volume and sensory responses to single-train rapid magnetic stimulation bilaterally on the neck (RMS_BAMPS_) in which the tidal volume achieved was between 4-8ml/kg of body weight. All dots represent a single data point. Black, white and grey dots represent RMS_BAMPS_ with the parabolic, D-shape and butterfly coil, respectively. Data points are not present in participant 1 and 10 as all tidal volume responses were outside of the 4-8ml/kg body weight range (both higher and lower values achieved for both participants).

### RMS_BAMPS_-ventilation trial

Eleven participants underwent the RMS_BAMPS_-ventilation trial (Figure 6) with the D-shape coil that did not overheat. Nine completed the 10-min protocol, one completed 6min (stopped due to discomfort; 7points), while in one participant ventilation could not be achieved. Five participants were hyperventilated compared to resting breathing resulting in an overall slight hyperventilatory response (P_ET_CO_2_=33.9±5.0 vs. 42.0±3.4mmHg, *P*=0.001; S_P_O_2_ 98±1%) caused by an increase in V_T_ (1.14±0.30 vs. 0.84±0.13L, *P*=0.008), but not breathing frequency (13.9±2.1 vs. 12.5±3.0 breaths per minute, *P*=0.105). Both 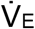 (*P*=0.156) and V_T_ (*P*=0.216) stayed constant over time. Mean SBP (141±9 vs. 122±10mmHg, *P=*0.003) and DBP (82±6 vs. 72±5mmHg, *P*=0.007) were elevated compared to baseline, while only SBP significantly increased throughout the trial (*P*=0.024). HR remained at baseline levels (64±8 vs. 62±8 beats per minute, *P*=0.165) and did not significantly change over time (*P*=0.101). Pain (10-min average: 1.2±1.1points), discomfort (2.5±1.5points), paresthesia (1.8±1.7points) and air hunger (1.1±1.7points) did not significantly change over the 10-min duration (all *P*≥0.387). Participants with balloon catheters (n=5) had a P_di,mean_ of 9.4±3.5cmH2O and a TTI_di,mean_ of 0.02±0.01 throughout the 10min, which did not change over time (*P*=0.833 and 0.937, respectively).

**Figure 6.**
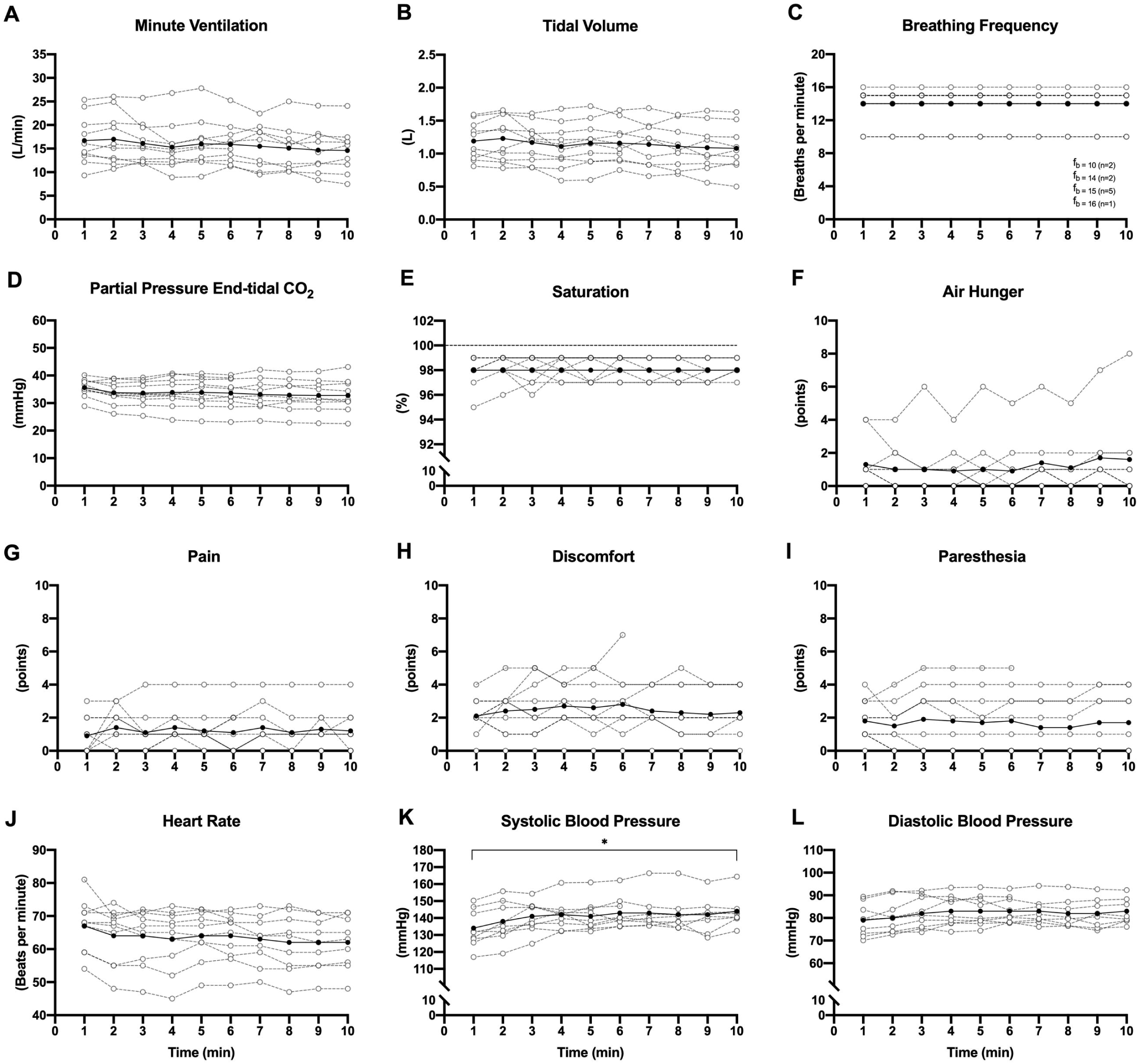
Ventilatory, sensory and cardiovascular responses to continuous rapid magnetic stimulation bilaterally on the neck. Black data points represent the group means for each 1-min block of ventilation, while white data points represent individual participants. Each participant was stimulated with the D-shape coil. *, significant regression.

### RMS_aMS_ on the chest

Eight participants underwent the single-train RMS_aMS_-protocol and only one participant (without balloons) showed a V_T_ response, which could only be achieved with the parabolic coil. Inspiration was achieved with all stimulation-frequencies, but required ≥40% of stimulator-output. The maximal tolerated stimulator-output was 50% (15, 20, 25Hz) and 40% (30Hz). The range of VT was 0.11-0.30L, while pain ranged between 5-7points, discomfort 7-8points and paresthesia 6-10points.

## Discussion

In the present study, RMS_BAMPS_ could elicit flow in ≈86% of participants with parabolic, D-shape and butterfly coils, and the resultant diaphragm contractions induced V_T_ similar or larger than during spontaneous resting breathing. The butterfly coil required the highest stimulator-output for V_T_ similar to the other coils which resulted in larger discomfort at maximal responses. In contrast, bilateral RMS_aMS_ could only elicit flow with the parabolic coil in one participant (13%). Finally, all but one participant (91%) reached 10min of continuous RMS_BAMPS_-induced ventilation, without coils overheating and without a decrease in 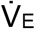 over time.

### Single-train RMS

While all three coils produced similar inspiratory responses, the butterfly coil required the largest stimulator-output despite having the largest maximal magnetic flux. The discrepancy between responses of different coils at the same stimulator-output likely reflects the butterfly being the largest coil making it difficult to optimally place often resulting in a small space between skin and coil surface. The higher stimulator-output needed to produce similar V_T_ also resulted in the largest discomfort at maximal responses, possibly due to the butterfly coil causing more dental pain.

Adler and co-workers (15) reported that 15Hz at 65% of stimulator-output served as the optimal setting using RMS_CMS_ to achieve the highest P_di,peak_ (≈20 cmH_2_O) with sensory ratings below 3points. Using the same criteria in the present study (pain below 3points; ≥4 participants equipped with balloons), the optimal settings would be 25Hz at 30% stimulator-output with the parabolic (P_di,peak_=14.8±4.5cmH_2_O), 20Hz at 40% with the D-shape (P_di,peak_=19.6±5.6cmH_2_O), and 15Hz at 40% with the butterfly (P_di,peak_=8.4±2.9cmH_2_O) coils. These P_di_, as well as maximal P_di_, are all lower than the ones of Adler *et al*. (15). However, this difference likely reflects that our participants kept their glottis open to allow airflow, while Adler *et al*.’s (15) kept their glottis closed during stimulation. Also, CMS is known to produce larger P_di_ twitches in response to single stimuli compared to bilateral stimulations (13, 21), attributed to recruitment of accessory respiratory muscles leading to chest wall stiffening (21), that likely also applies during RMS_CMS_. The diaphragm activation achieved in the present study in response to RMS_BAMPS_ reached and exceeded normal resting V_T_, and a level of stimulation likely able to attenuate diaphragm atrophy during MV. Recently, Sotak *et al*. (22) showed that percutaneous electrical PNS in ICU-patients during MV keeping the work of breathing between 0.2-2.0J/L reduced the rate of the development of diaphragmatic atrophy. However, in order to maximize therapeutic effects, methods to increase the level of activation without increasing V_T_ and thus without increasing the risk for lung injury, should be further explored.

Juxtapose to RMS_BAMPs_, RMS_aMS_ only induced inspiration in one participant. As such, the technique is likely not useful for diaphragm pacing. The difference between stimulation techniques likely reflects that RMS_aMS_ produced more movement of the shoulders, arms and torso compared to RMS_BAMPS_, which may have resulted in a shift in coil position (despite all efforts to avoid this) or an increased thickness of tissue between coils and phrenic nerves. The RMSaMS-responder was indeed small in stature (height=168cm, BMI=19.8kg/m^2^, female), but another like female (height=165cm, BMI=19.6kg/m^2^) did not show a flow response, possibly due to this participant’s lower tolerability of stimulator-output (40% vs. 60%). Also, flow could only be induced with the parabolic coil, possibly because this coil has a more focused point of stimulation compared to the others. In addition, V_T_ was lower, and all sensory ratings, especially paresthesia in the arms, were elevated compared to the mean responses of all participants at matched stimulation settings during single-train RMS_BAMPS_ (a protocol this participant did not perform).

Finally, a non-invasive abdominal belt may serve as a useful tool to quantify the amount of diaphragm activation during RMS given that Δ_Abdominal_ correlated with both V_T_ and P_di,mean_. Similarly, given that ΔGSR correlated with both pain and discomfort, it may serve as a useful measure of distress in unconscious patients (Table 3).

**Table 3.** Group and individual correlations for respiratory belts with inspiratory responses, and change in galvanic skin response with sensory responses during single-train rapid magnetic stimulation bilaterally on the neck.

| Correlated Variables | Group Repeated Measures Correlation |  | Individual Correlations |  |
| --- | --- | --- | --- | --- |
|  | r | P | Number of individuals with a significant correlation | Range of r (significant correlations only) |
| **ΔAbdominal Circumference** |  |  |  |  |
| ΔAbdominal and V<sub>T</sub> | 0.64 | <0.001 | 11 of 12 | 0.53 – 0.91 |
| ΔAbdominal and P<sub>di,mean</sub> | 0.76 | <0.001 | 6 of 6 | 0.69 – 0.93 |
| **ΔChest Circumference** |  |  |  |  |
| ΔChest and V<sub>T</sub> | 0.46 | <0.001 | 9 of 12 | 0.39 – 0.84 |
| ΔChest and P<sub>di,mean</sub> | 0.11 | 0.143 | 2 of 6 | -0.63 – 0.63 |
| **Galvanic Skin Response** |  |  |  |  |
| ΔGSR and Pain | 0.55 | <0.001 | 8 of 10 | 0.56 – 0.75 |
| ΔGSR and Discomfort | 0.63 | <0.001 | 9 of 10 | 0.52 – 0.87 |
| ΔGSR and Paresthesia | 0.38 | <0.001 | 5 of 10 | 0.39 – 0.78 |

*Definition of abbreviations:* ΔAbdominal = change in abdominal circumference; V<sub>T</sub> = tidal volume; P<sub>di,mean</sub> = mean transdiaphragmatic pressure; ΔChest = change in chest circumference; ΔGSR = change in galvanic skin response. Individual Pearson correlations were performed between respiratory belts and inspiratory variables, and individual Spearman correlations were performed between ΔGSR and sensory responses.

### Upper airway collapse

Similar to Sander *et al*. (14) (100%), RMS_BAMPS_ successfully induced flow in ≈86% of supine participants. In contrast, Adler *et al*. (15) were unsuccessful in inducing flow during RMS_CMS_ in sitting participants when instructed to keep their glottis open, which they attributed to UAC. Stimulation technique and the differences in neck position (flexed vs. extended - known to affect upper airway dynamics and flow pattern during phrenic nerve stimulation (23)) may contribute to this discrepancy. Notably, despite flow being induced in the majority of our participants, UAC was still prominent. All but one participant experienced full or partial UAC at least once during single-train RMS_BAMPS_ and two participants appeared to experience total UAC during all single-trains over two visits despite adjustments in stimulator-output, stimulation-frequency and coil positions. However, UAC may be an irrelevant issue in intubated and non-invasively ventilated patients given that Adler *et al*. (15) successfully alleviated UAC during RMS_CMS_ when positive pressure ventilation was added. Thus, positive airway pressure may be needed to avoid closure, but this needs to be explored more systematically.

### RMS_BAMPS_-ventilation

Continuous RMS_BAMPS_ induced ventilation in ten of 11 participants, while one participant experienced consistent UAC, despite responding to single-train RMS_BAMPS_. The 10-min limit was a result of our protocol rather than equipment overheating or participant intolerance (all that achieved 10min indicated they could go longer). Thus, with proper coil cooling, it is possible to overcome the 5-min limitation seen by Sander *et al*. (14).

Inducing ventilation via RMS_BAMPS_ appears to be a tolerable technique given the majority (90%) of participants with continuous ventilation were able to complete the 10-min protocol. In fact, mean perception of pain, discomfort and paresthesia were rated below 3points, however, one participant ended the trial early citing discomfort resulting from excessive shoulder movement on one side. This resulted from the coil moving out of the optimal position during the trial and reflects the importance of maintaining the same position between consecutive stimulations. All but two participants required slightly higher than 1-s stimulation duration (up to 1.3s) for optimal comfort, and six participants required a higher respiratory frequency compared to their resting breathing to suppress their urge to breathe. These adjustments occurred during familiarization based on their feedback and resulted in a slight hyperventilation in five cases. Perception of air hunger was rated ≤2 points at all times in all but two participants. It should be noted, however, that a brief break was taken between each minute to assess participant’s sensory ratings in which they resumed spontaneous breathing which may have contributed to reduced sensory ratings. Thus, the efficacy of ventilation without breaks and a longer than 10min still needs further exploration.

The mean 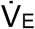 achieved in the present study was similar to the 14.0L/min by Sander *et al*. (14) with 25Hz at 40% stimulator-output (two MagStim butterfly coils) in 10 healthy volunteers. Their ventilation, however, surpassed the present when stimulator-output was increased to 50% (18.6L/min), but this increase in stimulator-output resulted in only three participants undergoing ventilation due to a largely increased perception of pain and discomfort, similar to our single-train RMS_BAMPS_ responses with maximal stimulator-output. The present study optimized RMS settings to each participant and was 20 (n=3), 25 (n=5) or 30 (n=2) Hz with a mean stimulator-output of 27% (range=20-40%). Stimulation parameters and thus V_T_ likely exceeded the levels that would be used or needed in mechanically ventilated patients in order to prevent lung injury. In any case, it is encouraging to note that in the present study with an excessive ventilation present, mean TTI_di_ was only 0.02 which is well below the 0.15 fatigue-inducing threshold (24). As such, RMS_BAMPS_ seems unlikely to cause fatigue-inducing contractions, at least in our group of young healthy participants, and as previously shown with CMS (15). To which extent muscle fatigue could play a role in various patient groups needs, however, further exploration.

Finally, while Sander *et al*. (14) reported no cardiovascular changes (unspecified), the present study showed an increased BP during RMS_BAMPS_-ventilation, but without changes in HR. Although the increase in BP did not reach an unsafe level in these young subjects and stimulator-output was likely higher than what would be used in patients, it is still suggested that the cardiovascular system is closely monitored during continuous RMS.

### Limitations

A few limitations remain. First, although flow traces were monitored to guarantee participants did not initiate inspiration, volitional assistance cannot be excluded with certainty. Second, the COVID-19 pandemic impacted the number of participants that returned for visit 2 and the optional visit. Third, not all participants tolerated balloon catheters resulting in a reduced sample with Pdi data, two being non-responders.

## Conclusion

RMS_BAMPS_, but not RMS_aMS_, can induce strong enough diaphragmatic contractions to ventilate healthy humans for 10min. However, we currently do not recommend replacing MV with RMS, but rather to use RMS to assist MV in order to potentially reduce ventilator-induced diaphragm atrophy. For most effective clinical use, newly designed equipment should be less bulky and optimized for PNS with stimulators automatically adjusting timing and output according to feedback from ventilation and non-desired side-effects.

## Data Availability

All data produced in the present study are available upon reasonable request to the authors.

## Acknowledgements

We would like to acknowledge the time, effort and tolerance of our participants during this difficult period of the COVID-19-pandemic, Prof. Dominique De Quervain (University of Basel, Switzerland) for providing the Brainsight Unit, Dr. Raniero Pittini and his team (Switzerland Innovation Park Biel/Bienne) for construction of the coil-fixation unit and providing Brainsight feedback instructions, Prof. Thomas Niederhauser and his team (Bern University of Applied Sciences, Switzerland) for valuable discussions and support, Daniel Leemann (Neurolite AG, Switzerland) for technical support, STIMIT AG (Switzerland) for technical, logistical and financial support, as well as innosuisse (34221.1 IP-LS) for financial support.

